# Effect of various hormonal and non-hormonal contraceptive methods on the vaginal milieu -A narrative review

**DOI:** 10.1101/2022.05.17.22275192

**Authors:** Deepti Tandon, Anushree D Patil, Mayuri Goriwale

**Affiliations:** Department of Clinical Research, ICMR- National Institute for Research in Reproductive and Child Health (NIRRCH), J.M. Street, Parel, Mumbai 400012

**Keywords:** Contraception OR Nugent score, Contraception OR Vaginal microbiome, Contraception OR Bacterial vaginosis, Contraception OR Vaginal pH

## Abstract

Globally the use of modern contraceptive methods has risen to 851 million. Use of these contraceptive methods does influence the vaginal health, which in turn affects the susceptibility towards acquiring RTI/STI. Methods to assess the vaginal health have also evolved over decades. The objective of this narrative review is to assess the influence of contraceptive methods on the vaginal health and also evaluate the methods used to assess the vaginal milieu. Suitable articles published in literature from 2007-2020 were identified from PubMed, Google Scholar using relevant keywords. Hormonal contraceptives included were combined oral contraceptive pills (COCP), Depot medroxyprogesterone acetate (DMPA)and Levonorgestrel IUCD(LNG-IUS). Non hormonal methods included were barrier methods, copper IUCD, diaphragm and vaginal sponge. Outcome parameter recorded were Nugent score, vaginal pH, bacterial vaginosis or defined microbiome profile.

COCP have been shown to protect the vaginal ecosystem primarily due to their oestrogen component. The use of IUCD causes initial dysbiosis chiefly due to associated irregular bleeding but long term use of LNG IUS stabilizes the microbiome. Use of injectable DMPA though does not increase the susceptibility to HIV but can promote growth of anaerobic organisms. Literature regarding condom, diaphragm and sponge is very scanty to draw a meaningful conclusion. Hence contraceptive methods can affect the vaginal health. There is need to periodically assess the vaginal milieu using test which is appropriate as per available expertise, infrastructure and cost and treat vaginal dysbiosis in respective cohorts to prevent reproductive morbidity.

## 1. Introduction

Worldwide as per world family planning highlights 2020, the number of women using modern or traditional methods of contraception are 851 and 85 million respectively.^1^ The vaginal ecosystem is dynamic, vulnerable and susceptible to change in response to exposure to various external stimuli such as hormones and allergens. ^2^Any positive or negative changes in these vaginal health is termed as vaginal eubiosis or vaginal dysbiosis respectively. This vaginal eubiosis is protective but the dysbiosis makes the women prone to various degrees of common but significant reproductive morbidities such as bacterial vaginosis, recurrent vaginal infections, risk of acquiring sexually transmitted infections, urinary tract infection, HIV transmission, pelvic inflammation and preterm births.^3,4^ There have been conflicting evidence in literature regarding the effect of available contraception methods on the vaginal health of women. The impact of these hormonal and non-hormonal contraceptives is dependent on the host factors which are further affected by the local immune responses, vaginal bacterial communities, competition and release of bactericidal products, or any inflammation acquired due to prolonged use.^5^

This narrative review will help us understand the effect of various hormonal and non-hormonal contraceptive methods on vaginal health and further evaluate the various methods used to assess vaginal milieu

## 2. Methodology

In this narrative review the search engines that were explored were Pubmed and Google Scholar. The following keywords were used for the research: Contraception and Vaginal microbiome OR Contraception and Nugent score OR Contraception and Bacterial vaginosis OR Contraception and vaginal pH. The studies which met the following criteria were included in the current review

1. Prospective studies published in English

2. Published in last fourteen years 2007-2020 irrespective of data collection period

3. Studies mentioning the contraceptive method used

## 3. Results

The studies and their findings are described in brief. Data is compiled and interpreted to give a comprehensive overview of effect of various contraception method on the vaginal milieu.

Individual contraceptives are discussed under the following subheading-

### 3.1. IUCD and vaginal milieu

Copper containing devices such as copper T 380 A and copper T 375 are more commonly used and provide contraceptive benefits up to ten years and five years respectively. Hormonal IUCD contain levonorgestrel in variable quantities and have effectiveness between 3-5 years. Commonly used hormonal IUCD are Mirena, Skyla, Liletta and Kyleena.

As reported in literature various studies have been done to evaluate the effect of copper T as well as LNG IUCD on the vaginal health of women.^6,7,8,9,10,11,12,13,14,15,16^ These studies have been enumerated in Table 1.

**Table I:** Studies evaluating effect of Intrauterine contraceptive devices on vaginal milieu.

| Sr No | Author Year | Study Setting | Sample Size | Contraceptive Evaluated | Test used | Result |
| --- | --- | --- | --- | --- | --- | --- |
| 1. | Giraldo et al 2019<sup>6</sup> | Brasil | 60 | LNG IUCD<sup>*</sup> | pH, Pap smear, Gram Stain | No changes in vaginal pH, vaginal candidiasis, BV<sup>□</sup>, vaginal microbiota |
| 2 | Achilles et al 2018<sup>7</sup> | Zimbabwe | 93 | Copper T | PCR | Mean increase in Nugent score was 1.2 |
| 3 | Sharma et al 2018<sup>8</sup> | Toronto | 17 | Copper T<br>LNG IUCD<sup>*</sup> | Cytokine | Increase in inflammatory cytokines |
| 4 | Brooks et al 2017<sup>9</sup> | USA | 682 | LNG IUCD<sup>*</sup> | 16S rRNA sequencing | *Prevotella*, *Sneathia*, *Dialister*, and *Megasaera* species were significantly more abundant in LNG-IUS users |
| 5 | Bassis et al 2017<sup>10</sup> | USA | 76 | Copper T<br>LNG IUCD<sup>*</sup> | 16S rRNA sequencing | No evidence of altered vaginal microbiota composition. |
| 6 | Donders et al 2017<sup>11</sup> | Belgian | 252 | LNG IUCD<sup>*</sup> | Gram stain, Culture | Initial three months increase BV, later stabilizes. More vaginal colonization with *Candida* |
| 7. | Jacobson et al 2014<sup>12</sup> | - | 11 | LNG IUCD<sup>*</sup> | 16S rRNA sequencing | No change in vaginal microbiome |
| 8 | Madden et al 2012<sup>13</sup> | - | 153 | Copper T | Gram stain | Intermediate flora, and irregular vaginal bleeding were significantly associated with BV<sup>□</sup> |
| 9 | Teeraganok | Thailand | 92 | Copper T | Amsel's | The prevalence of BV<sup>□</sup> was higher |

|  | et al 2012<sup>14</sup> |  |  | Non users | criteria | among copper IUD<sup>††</sup> group |
| --- | --- | --- | --- | --- | --- | --- |
| 10 | Neale et al<br>2009<sup>15</sup> | Truro | 172 | Copper T | Symptoms | More likely to have developed an abnormal vaginal discharge 4–6 weeks after insertion |
|  |  |  |  | LNG IUCD<sup>*</sup> |  |  |
| 11 | Lessard et al 2007<sup>16</sup> | Brazil | 187 | LNG IUCD<sup>*</sup> | Cytology | Increased frequency of candida but no cytological changes after seven years |

<sup>\*</sup> LNG IUCD Levonorgestrel Intra-uterine Contraceptive Device <sup>†</sup> BV- Bacterial vaginosis, <sup>††</sup> IUD-Intrauterine device

As inference from Table 1, literature does suggest an association between use of copper IUCD and dysbiotic vaginal health leading to increased vaginal discharge, bacterial vaginosis, increase in inflammatory cytokines and increased colonization by candida.^7,8,11,12,13,14,15^ Further evaluation of the effect of levonorgestrel intrauterine system (LNG-IUS) on lower genital tract is conflicting. Literature does suggest that there is increase in prevalence of dysbiotic microbiome profile and raised cytokine profile after use of LNG IUCD. ^8,9^There has been an observation of increased colonization of candida species after long term use as suggested by Donders et al and Lessard T et al. Reactional changes in the vaginal environment have also been observed in a recent study by Giraldo et al.^6,11,16^ Few studies have reported a positive influence on the community stability with use of LNG IUCD but studies by Bassis et al and Jacobson et al found no evidence that LNG contraception altered the vaginal microbiota composition.^10,12,17^

### 3.2. Oral contraceptives and vaginal milieu

Oral contraceptives are hormonal contraceptives containing oestrogen and progesterone or only progesterone. The most commonly used is the one containing both oestrogen and progesterone.

Fosch et al in 2017 used the Balance of Vaginal Content (BAVACO) methodology to assess the vaginal health after use of this contraceptive. This methodology used two parameters-Nugent scoring and vaginal inflammatory reaction to give a composite score of vaginal health.^18^ Continuous and sustained use of oral contraceptive pills over a period of six months led to the stability of vaginal ecosystem. *Lactobacillus crispatus* followed by *Lactobacillus jensenii* has been found to be a predominant organism by Brooks et al and Fosch et al after use of combined oral contraceptive pills which in turn was protective species and promoter of vaginal health.^9,18^Studies as summarized in table 2 shows the various studies in literature which have evaluated the use of oral contraceptive pills and on vaginal health.^18,19,20,21^ Majority of the studies show a relatively stable vaginal ecosystem after the use of oral contraceptive pills. Brotman et al and Rifkin et al showed stable bacterial communities in various cohorts respectively.^19,20^ Increased oestrogen though promoted the vaginal colonization by yeast cells in oral contraceptive users.^18^

**Table II:**
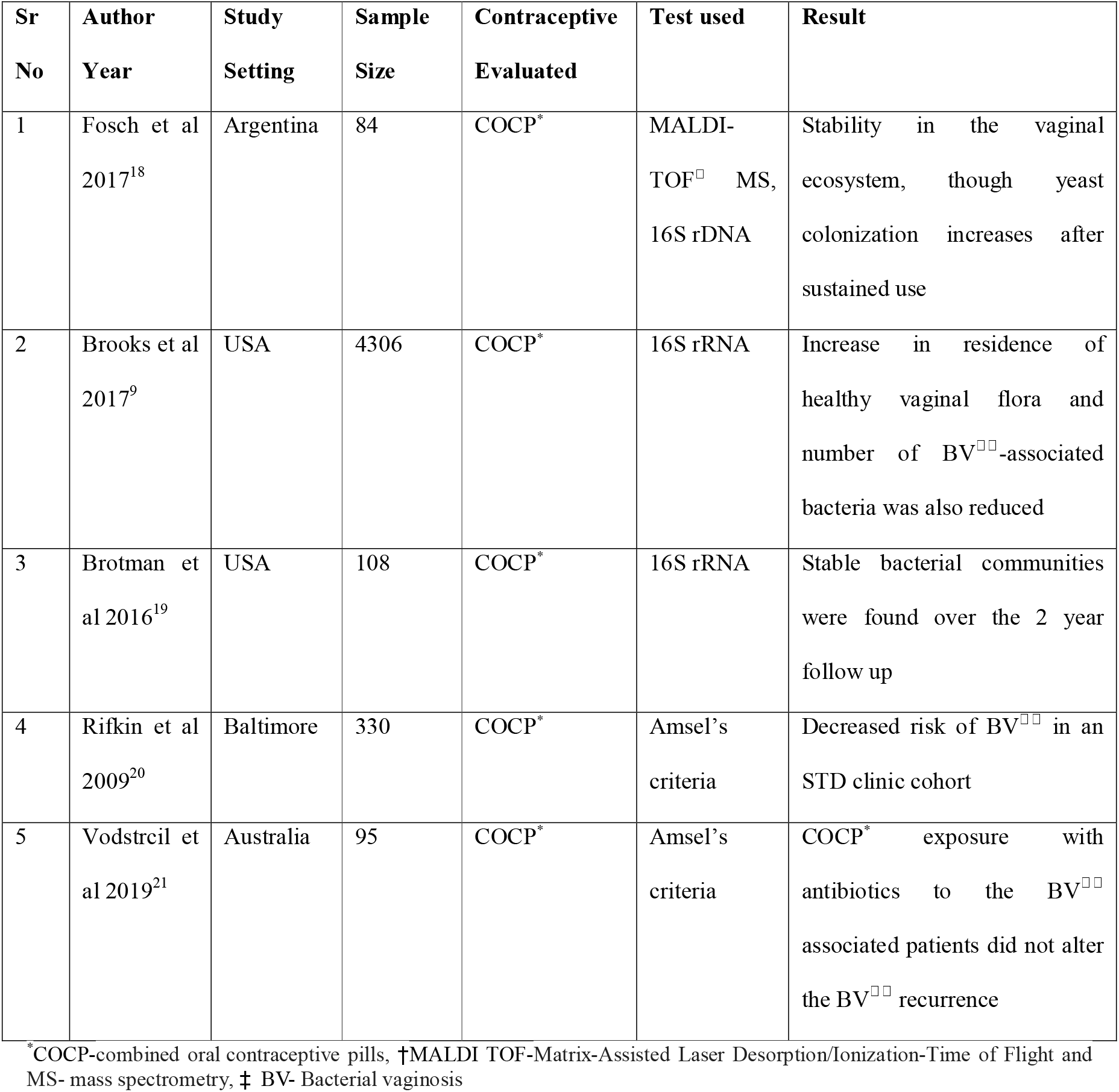
Studies evaluating effect of Combined oral contraceptive pills on vaginal milieu.

### 3.3. DMPA and vaginal milieu

Depo-Provera or Depot medroxyprogesterone acetate is progesterone containing injectable contraceptive, which is administered every after three months.

Meta-analysis done by Ralph et al echoed that there was moderate increase in HIV acquisition especially in high-risk population after DMPA use.^22^ Yang et al in 2019 pointed out that the microbiome becomes diversified and shows higher colonization of *Prevotella* OTUs after continuous use of DMPA, though it’s the racial diversity in the vaginal microbiome which determines their susceptibility towards acquiring STI.^23^

Retrospective study by Brooks et al in 2017 suggest that use of DMPA is associated with increase in colonization by *A. vaginae* and *P. Biviano*, hence a detrimental effect was seen on the vaginal microbiota.^9^ Animal studies on pigtail macaques by Butler K also validated that risk of HIV acquisition is increased in a dose dependent manner.^24^ Though there was mounting evidence that the risk of STI especially HIV transmission is increased with use of DMPA, the result of ECHO Study (Evidence for Contraceptive Options in HIV Outcomes) published on June 13, 2019 shows that there was no risk of acquiring HIV infection. This largest multicentre, open label, randomized trial done between 2015 to 2017 across four centres on 2609 women using DMPA suggest no substantial risk of HIV.^25^ Hence summarization of all these studies as tabulated in Table 3 suggest that the vaginal microbiota is probably altered with increase in the species especially *Prevotella* OTUs and *A. vaginae*. ^26,27,28^

**Table III:**
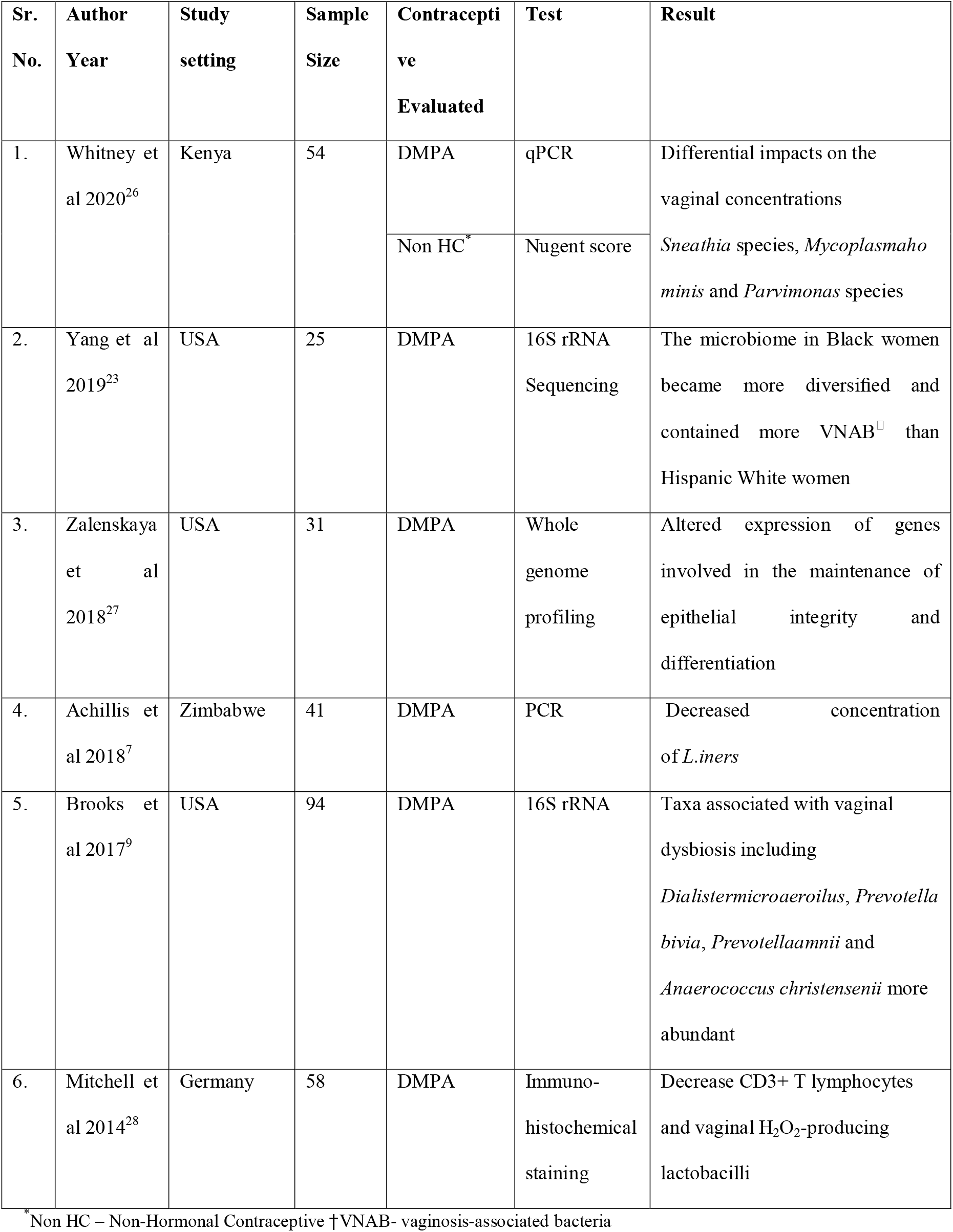
Studies evaluating effect of Depot medroxyprogesterone acetate(DMPA) on vaginal milieu.

### 3.4. Condom and Vaginal Milieu

Condoms are one of the non-hormonal reversible, cost effective and easy to use contraception method used by many men during the intercourse. Use of condoms has prevented the transmission of Sexually Transmitted Infections (STIs) worldwide.

Fosch et al. in 2017 studied the vaginal ecosystem of eight women after continuous use of condom. As per the study, there was an increase in the vaginal inflammatory reaction due to latex allergy. *L. Gasseri* was the most commonly found followed by *L. crispatus, L. jensenii, L. murinus* and *L. Johnsonii* in the order.^*18*^

Liyan Ma et al. in 2013 studied effect of use of condom on the vaginal milieu. The study concluded that consistent effect of condoms increases the colonization of *Lactobacillus crispatus* which indeed was protective for bacterial vaginosis and HIV.^29^ Table 4 elaborates the studies of vaginal mileu after condom use.

**Table IV:** Studies evaluating the effect of condom on vaginal milieu.

| **Sr. No.** | **Author Year** | **Study setting** | **Sample Size** | **Contraceptive Evaluated** | **Test used** | **Result** |
| --- | --- | --- | --- | --- | --- | --- |
| 1 | Fosch et al 2017<sup>9</sup> | Argentina | 8 | Condom | Culture,16S rRNA sequencing | Increase in the vaginal inflammatory reaction due to latex |
| 2 | Liyan Ma et al 2013<sup>29</sup> | Canada | 164 | Condom | polymerase chain reaction(PCR), Nugent score & quantitative culture | *Lactobacilli crispatus* had higher colonization which was protective for BV and HIV |

### 3.5. Miscellaneous contraception

Some other contraception methods that are evaluated to check the effect on vaginal ecology are Nonoxynol-9, Progesterone vaginal rings and latex diaphragm.^30^ Use of Nonoxynol 9 vaginal sponge had a dose dependent effect on increasing the colonization of anaerobic Gram negative rods, but the use of progesterone ring improved the Nugent score.^31^ The use of diaphragm did not demonstrate increased disruption of vaginal ecosystem.^32^

### 3.6. Methods to assess vaginal health

Various tests have evolved over past decades to assess the vaginal health. Vaginal pH is measured using vaginal pH strips and gloves, have high sensitivity in the range 72-79 %, but low specificity of 60-53% respectively.^33^ Cervicovaginal lavage for cytokine analysis and vaginal epithelial exfoliation measurement are also used to assess the vaginal health.^8,34^ These specimen can be used to measure inflammatory markers like cytokines, free glycogen levels and exfoliated epithelial cells levels of which are associated with growth of different Lactobacillus species.^35^ Further culturing techniques have allowed more specific strains to be identified, but vaginal microbiome analysis has gone step ahead to allow taxonomic and metabolomics analysis of the bacterial communities with respect to various vaginal infections. Two advanced microbial and metabolome techniques of next generation sequencing (NGS) and proton-based nuclear magnetic resonance spectroscopy are being used to identify the microbial picture and identify the functional correlation to various vaginal morbidities by analysing the metabolities.^36^

## 4. Discussion

It seems worthwhile to assess the literature related to vaginal milieu in women using various contraceptives to understand the short- and long-term effects of these contraceptives with respect to evolving literature of their effect on vaginal heath.

The use of IUCD does seem to negatively affect the vaginal health during the initial period of use, probably because of the associated menstrual disturbances. Madden et al explained this association in 2012 possibly due to irregular vaginal bleeding which is commonly found in IUCD users. This blood increases the acidic pH of vagina as well as causes the agglutination of lactobacillus to red blood cells which may result in a decreased vaginal *Lactobacillus spp*. concentration in women with frequent or persistent uterine bleeding.^13^ The association between LNG IUCD and initial dysbiosis can be explained by the fact that use of LNG IUCD is associated with irregular vaginal bleeding initially and later there is cessation of blood flow after prolonged use, hence the microbiome which destabilizes after initial use and later balances itself. Donders et al study in the 2018 stated that, symptomatic or asymptomatic *Candida* colonization runs parallel to the growth of *Lactobacillus* because of preferential colonization of *Candida* with *Lactobacilli*.^11^ Long-term use of LNG IUCD stabilizes the microbiome because of improvement in menstrual cycle. However more prospective and functional studies are required with use of LNG IUCD.

Two mechanisms are identified as how oestrogen and progesterone present in oral pills contribute to vaginal health. Hormonal stimulation leads to predominance of epithelial and intermediate cells and lysis of the latter leads to release of glycogen, which promotes the growth of *Lactobacillus spp*. like *L*.*crispatus* and *L*.*Gasseri*. These species coverts glycogen into lactic acid and H_2_O_2,_ which helps in improvising vaginal health by acting as bactericides. Activity of proton pump in the vaginal and cervical epithelial cells is enhanced in the presence of oestrogen hormone thereby decreasing the vaginal pH.^7,37^ Norgestrel, levonorgestrel and desogestrel are commonly used as a second and third generation progesterone in oral contraceptive formulation.As highlighted by Achilles et al, the effect of progesterone on vaginal health will depend on their affinity to attach to the glucocorticoid receptors, mineralocorticoid receptor and androgen receptor, which are present in various cell. Activation of these leads to immunosuppression and mediates the non-contraceptive biological effects. The progesterone used in current OC formulation seem to have less effect on the glucocorticoid and mineralocorticoids receptor which in turn lowers the chances immune and genital infection susceptibility.^7,18^Medroxyprogesterone acetate and gestodene are the only progesterone that have high affinity for this receptor. The rest have low affinity thereby less potential to effect the vaginal health.

DMPA is a progesterone with significant immunosuppressive effects as it strongly binds to the glucocorticoid receptors. Use of DMPA was thought to have a potential association between reproductive tract infection especially HIV transmission because of the hypoestrogenic state caused by prolonged use of DMPA leading to down regulates various genes involved in the maintenance of mucosal integrity.^27,38^ Though use of DMPA does not increase the risk of HIV infection as shown by ECHO trial, there is still evidence of increase in the colonization of anaerobic bacteria especially Prevotella OTUs and *A. vaginae* which further increase the potential for acquiring bacterial vaginosis.^25^ This effect can also still be explained due to the prolonged hypoestrogenic state which decreases the glycogen in cells hence decrease substrate for Lactobacillus along with associated disturbed menstrual pattern promotes growth of anaerobic species in DMPA users. Romas et al in a recent study has suggested that DMPA causes increased cytokine profile, epithelial thinning and immunosuppressive response .^39^

Though it is well known that use of condom is protective against sexually transmitted infections, less literature is available on the specific vaginal changes and microbiome associated with condom use. However, few studies states condom use along with dominance of *Lactobacillus crispatus* is protective against sexually transmitted infections and HIV by enhancing the entrapment of these virus in the cervicovaginal mucosa.^40^ This species of *Lactobacillus crispatus* has been shown to be protective against Chlamydia trachomatis, Gardenella vaginalis and Neisseria gonorrhoea infections too. However, the effect of other non-hormonal method on the vaginal health remains inconclusive owing to scanty literature available in the context. The peculiar microbiome associated with these contraceptive methods are not well explored and far less studies are available.

Evaluation of methods to assess vaginal health show that Vaginal pH is a relatively simple tool, but has disadvantages of false reading due to presence of mucus and semen inside the vagina.^33^ Amsel’s criteria described by Amsel in 1983 is not a gold standard method to assess the vaginal health owing to the component of whiff test which is very subjective but this test is favoured by clinicians as it eliminates the need of cumbersome laboratory equipment. The Amsel’s criteria however does require some skill in microscopy to identify the clue cells and both conventional and modified Amsel’s criteria hold similar diagnostic accuracy.^41,42,43^Nugent score still remains the benchmark test to assess the vaginal health. Additional methods as illustrated by Sharma et al in 2018 who measured the cervico-vaginal cytokine levels highlighted the importance of cellular and cytokine milieu of the female genital tract mucosa which in turn determines the susceptibility for sexually tract infections. Non culture-based modern technologies like 16S rRNA gene sequencing greatly facilitate comprehensive surveys of the composition of vaginal microbial communities in comparison with the conventional methods. These molecular methods hold better speed, accuracy as well as ability to identify plethora of species which may not grow on culture media. Use of these advanced technologies has helped in taxonomic and metabolomics analysis of the vaginal ecosystem thereby helping to characterize the peculiar microbiome and metabolite associated with particular vaginal infection. This in turn may be useful in future to improve the therapeutic interventions. Exploratory studies to evaluate the therapeutic options of vaginal microbiome transplantation in targeted subpopulations in underway. But the cost factor and elaborate infrastructure attached to it makes it of limited benefit in our day-to-day clinical work.

## 5. Conclusion

The use of various available contraceptives is absolutely necessary but it does effect the vaginal health and further reproductive health of a women. There is need to periodically assess the vaginal milieu using various tests, which are appropriate as per available expertise, infrastructure and cost. Any dysbiosis should be immediately treated to prevent reproductive morbidities.

## Data Availability

All data produced in the present work are contained in the manuscript

## Acknowledgements

The authors acknowledge the encouragement and guidance received from Director, ICMR-NIRRH and Dr Sanjay Chauhan Head of Operational & Clinical research ICMR NIRRH.

